# A Population Study of Clinical Trial Accrual for Women and Minorities in Neuro-Oncology Following the NIH Revitalization Act

**DOI:** 10.1101/2021.05.28.21258034

**Authors:** Sheantel J. Reihl, Nirav Patil, Ramin A. Morshed, Mulki Mehari, Alexander Aabedi, Ugonma N. Chukwueke, Alyx B. Porter, Valy Fontil, Gino Cioffi, Kristin Waite, Carol Kruchko, Quinn Ostrom, Jill Barnholtz-Sloan, Shawn L. Hervey-Jumper

**Author notes:** Authorship: Research Concept and Plan: SJR, UNC, ABP, VF, SLHJ. Data collection & analysis: SJR, NP, RAM, MM, GC, KM, CK, QO, JBS, SLHJ. Manuscript preparation: all authors. Corresponding Author: Shawn Hervey-Jumper. **Funding** National Institute of Health, Award Number: TL1TR001871-05 (SR). National Center for Advancing Translational Sciences of the NIH (RAM), Neurosurgery Research and Education Foundation (RAM), Robert Wood Johnson Foundation 74259 (SHJ), NINDS K08 110919-01 (SHJ), Loglio Collective (SHJ). The CBTRUS data were provided through an agreement with the Centers for Disease Control’s National Program of Cancer Registries. In addition, CBTRUS used data from the research data files of the National Cancer Institute’s (NCI) Surveillance, Epidemiology, and End Results Program, and the National Center for Health Statistics National Vital Statistics System. CBTRUS acknowledges and appreciates these contributions to this report and to cancer surveillance in general. Funding for CBTRUS was provided by the Centers for Disease Control and Prevention (CDC) under Contract No. 75D30119C06056, the American Brain Tumor Association, The Sontag Foundation, Novocure, the Musella Foundation, National Brain Tumor Society, the Pediatric Brain Tumor Foundation, the Uncle Kory Foundation, the Zelda Dorin Tetenbaum Memorial Fund, as well as private and in-kind donations. Contents are solely the responsibility of the authors and do not necessarily represent the official views of the CDC or the NCI.

## Abstract

**BACKGROUND:** The NIH Revitalization Act, implemented 29 years ago, set to improve the representation of women and minorities in clinical trials. In this study, we investigate progress made in all phase therapeutic clinical trials for neuro-epithelial CNS tumors stratified by demographic-specific age-adjusted disease incidence and mortality. Additionally, we identify workforce characteristics associated with clinical trials meeting established accrual benchmarks.

**METHODS:** Registry study of published clinical trials for World Health Organization defined neuro-epithelial CNS tumors between January 2000 and December 2019. Study participants were obtained from PubMed and ClinicalTrials.gov. Population-based data originated from the CBTRUS for incidence analyses. SEER-18 Incidence-Based Mortality data was used for mortality analysis. Descriptive statistics, Fisher exact, and χ^2^ tests were used for data analysis.

**RESULTS:** Among 662 published clinical trials representing 49,907 participants, 62.5% of study participants were men and 37.5% were women (P<0.0001) representing a mortality specific over-accrual for men (P=0.001). Whites, Asians, Blacks, and Hispanics represented 91.7%, 1.5%, 2.6%, and 1.7% of trial participants. Compared with mortality, Blacks (47% of expected mortality, P=0.008), Hispanics (17% of expected mortality, P<0.001) and Asians (33% of expected mortality, P<.001) were underrepresented compared with Whites (114% of expected mortality, P<0.001). Clinical trials meeting accrual benchmarks for race included minority authorship.

**CONCLUSIONS:** Following the Revitalization Act, minorities and women remain underrepresented when compared with their demographic-specific incidence and mortality in therapeutic clinical trials for neuroepithelial tumors. This study provides a framework for clinical trial accrual efforts and offers guidance regarding workforce considerations associated with enrollment of vulnerable patients.

**Key Points:** Minorities and women with brain tumor diagnosis remain significantly under-accrued for neuro-oncology clinical trials compared to Caucasians and men based on proportional disease burden and demographic-specific mortality.

**Importance of the Study:** The current state of clinical trial accrual in the U.S. for adult patients with gliomas across different demographic groups has not been comprehensively studied. This study aims to quantify clinical trial accrual by age-adjusted disease incidence and mortality for gender and race during a 20-year period following the NIH Revitalization Act, and to identify workforce characteristics associated with clinical trials meeting established race and gender accrual benchmarks. Minorities and women with brain tumor diagnosis remain significantly under-accrued for neuro-oncology clinical trials compared to Caucasians and men based on proportional disease burden and demographic-specific mortality. Despite the enactment of the NIH Revitalization Act to improve the representation of women and minorities in clinical trials nearly 30 years ago, this goal remains unmet in the field of neuro-oncology. Significant work is required to continue to implement and improve interventions to increase accrual of diverse patient populations.

## Introduction

Patient specific factors such as race and gender remain essential contributors to an individual’s health and wellness in the United States (U.S.). National policies such as the 1993 National Institutes of Health (NIH) Revitalization Act established guidelines for the inclusion of women and minorities in clinical research. The statute outlined the necessary components in design, implementation, and outreach to include under-represented populations, consistent with their representation in the U.S. population (currently 51% women, 36.3% minorities)^1,2,3^. Additional guidance offered a framework for enrollment based on disease specific race and gender incidence.

Despite this legislation, over the last 30 years, in general clinical trial participants remain largely young, white, and male^4,5,6,7^. A review of clinical trials associated with U.S. Food and Drug Administration (FDA) approved cancer therapies from 2008-2018, found that 15 years following the NIH Revitalization Act, only 63% of trials reported race, and 7.8% reports the four major racial groups in the U.S.^8^ Furthermore, within FDA approved cancer trials, Black and Hispanic patients accounted for 3.1% and 6.1% of trial participants respectively, far below cancer incidence.

Neuro-epithelial brain tumors such as diffuse gliomas are the most common adult primary brain tumors in the United States, accounting for over 50% of malignant brain cancers. For these patients, many will exhaust standard of care treatment options allowing clinical trials to be widely accepted as the highest quality care, with participation being associated with improved clinical outcomes and increased survival^9^. Access to clinical trials not only allows for generalizability of scientific research, but it also provides for equitable treatment of diverse patient populations. Knowledge regarding clinical trial participation among women and minorities at the population level in neuro-oncology is a critical knowledge gap.

In neuro-oncology, the importance of race and gender diversity towards ensuring equity, validity, and generalized interpretability of results is of great importance. Our goal is to provide a framework for understanding clinical trial enrollment for brain cancer patients by (1) reviewing clinical trial gender and race reporting, (2) quantifying proportion of study participants stratified by age adjusted disease specific incidence and mortality, (3) use population data to determine how these findings have changed over a 20-year period following institution of the NIH Revitalization Act, and (4) identify workforce characteristics associated with optimal clinical trial accrual. This study will provide a framework for investigating clinical trial participation based on disease burden through direct evaluation of incidence and mortality rates.

## Methods

### Clinical Trial Accrual Data

Based on World Health Organization (WHO) 2016 diagnostic criteria, study enrollment included the following neuro-epithelial tumor diagnosis: diffuse astrocytoma, anaplastic astrocytoma, glioblastoma, oligodendroglioma, anaplastic oligodendroglioma, ependymal tumors, glioma malignant, not otherwise specified (NOS). A systematic review of the literature was conducted through a PubMed query to identify articles published of clinical (Phase I-IV) trials of adult gliomas between January 1, 2000 and December 31, 2019. Adult (18+) participants with a primary glioma diagnosis were included for analysis. For the studies conducted in the U.S., “Minority” status was defined as patients belonging to any of the NIH-designated race based underserved groups. This includes individuals with the following racial or ethnicity makeup: Asian/ Pacific Island native, African American/Black, Hispanic/Latinx, and Native American/Alaska Native. If participant demographics were not explicitly reported in the article, and the trial’s national clinical trial (NCT) number was available, a subsequent search was conducted on ClinicalTrials.gov.

### Incidence & Mortality Data

The following datasets were utilized and are described in detail below: *CBTRUS Incidence Data:* Central Brain Tumor Registry of the United States SEER*Stat Database. Centers for Disease control (CDC) National Program of Cancer Registries (NPCR) and National Cancer Institute (NCI) Surveillance, Epidemiology and End Results (SEER) Incidence Data, 2019 submission (2000-2017)^12^. *SEER Incidence-Based Mortality Data:* SEER Program (www.seer.cancer.gov) SEER*Stat Database: Incidence-Based Mortality - SEER Research Data, 18 Registries, Nov 2019 Sub (2000-2017) - Linked to County Attributes - Time Dependent (1990-2017) Income/Rurality, 1969-2018 Counties, National Cancer Institute, DCCPS, Surveillance Research Program. Released April 2020, based on the 2019 submission^15^.

The Central Brain Tumor Registry of the United States (CBTRUS) database (Data provided by CDC’s NPCR and NCI’s SEER Program, 2000-2017) was used to estimate the age-adjusted incidence rates. Average annual age-adjusted incidence rates and 95% confidence intervals were estimated per 100,000 population, based on one-year age groupings and standardized to the 2000 US standard population by race and ethnicity (Supplemental Table 2)^14^. Incidence-based age-adjusted mortality rates were calculated using the data from the SEER 18 (Supplemental Table 3) by race and ethnicity.

### Diversity Data Among High Accruing Programs

Clinical trials which reported minority recruitment at or above 10% of participants (equivalent to the upper tercile of the distribution) were identified. Descriptive statistics were reported from self-reported faculty diversity in the neurology, medical oncology, and neurosurgery departments, contrasted with 2018 Association of American Medical Colleges (AAMC) National Physician Workforce data along with Census Bureau 2010-2016 City Demographic data.

### Data Analysis

This study reports descriptive statistics of enrollment proportions for each demographic group for the 20-year study period, and year-over-year trends. Incidence and mortality counts, rates, and other relevant statistics were calculated using SEER*Stat 8.3.8^16^. Primary enrollment disparity is reported as the difference in proportions (DF) between accrual and mortality. A secondary comparison is reported in the supplemental data examining “accrual vs incidence”, by group. Z and χ^2^ tests of proportions were used to evaluate the significance of the associations in comparison groups and odds ratios are used to describe enrollment ratios. The level of significance was p<0.05 for all analyses. Group level statistics were performed using STATA SE 16.

## Results

### Clinical trial reporting accrual by demographic group

An initial search returned 1,932 articles that met inclusion criteria. After screening, 662 full text articles were identified that reported patient roster with demographic information of accrued participants. These 662 articles included 49,907 enrolled participants published during the 20-year period. Of these, 527 articles (including 41,933 participants) specifically reported the distribution of sex in the study. One hundred and thirty articles (including 11,943 participants) reported participants of White race, while 104 of those articles specified participants belonging to any minority racial or ethnic group (Supplemental Figure 1). Importantly, while 80% of eligible articles reported accrual by sex, only 20% of articles reported any racial breakdown, where many listed numbers of “White” participants only, and only 16% reported the number accrued of any minority race or ethnicity (Table 1).

**Table 1.** Participant Demographics Reported in Published Glioma Clinical Trial Articles 2000-2019. AA: African American, PI: Pacific Islander, AN: Alaska Native

|  |  | **N Articles**<br>**662** | **% Articles** | **N Patients**<br>**49,907** | **% Patients** | **% of Reported** |
| --- | --- | --- | --- | --- | --- | --- |
| Gender |  | 527 | 80% | 41,933 | 84% | - |
|  | Male |  |  | 26,237 | 53% | 63% |
|  | Female |  |  | 15,696 | 31% | 37% |
| Race |  | 130 | 20% | 11,943 | 24% | - |
|  | Minority (Any) | 104 | 16% | 663 | 1.3% | 5.6% |
|  | White | 130 | 20% | 10,806 | 22% | 90.5% |
|  | Black/AA | 66 | 10% | 256 | 0.5% | 2.1% |
|  | Asian/PI | 47 | 7% | 158 | 0.3% | 1.3% |
|  | American Indian/AN | 10 | 2% | 20 | 0.04% | 0.2% |
|  | Other/Unknown | 57 | 9% | 464 | 0.9% | 3.9% |
| Ethnicity |  |  |  |  |  |  |
|  | Hispanic/Latino | 36 | 5% | 239 | 0.5% | 2.0% |

**Figure 1.**
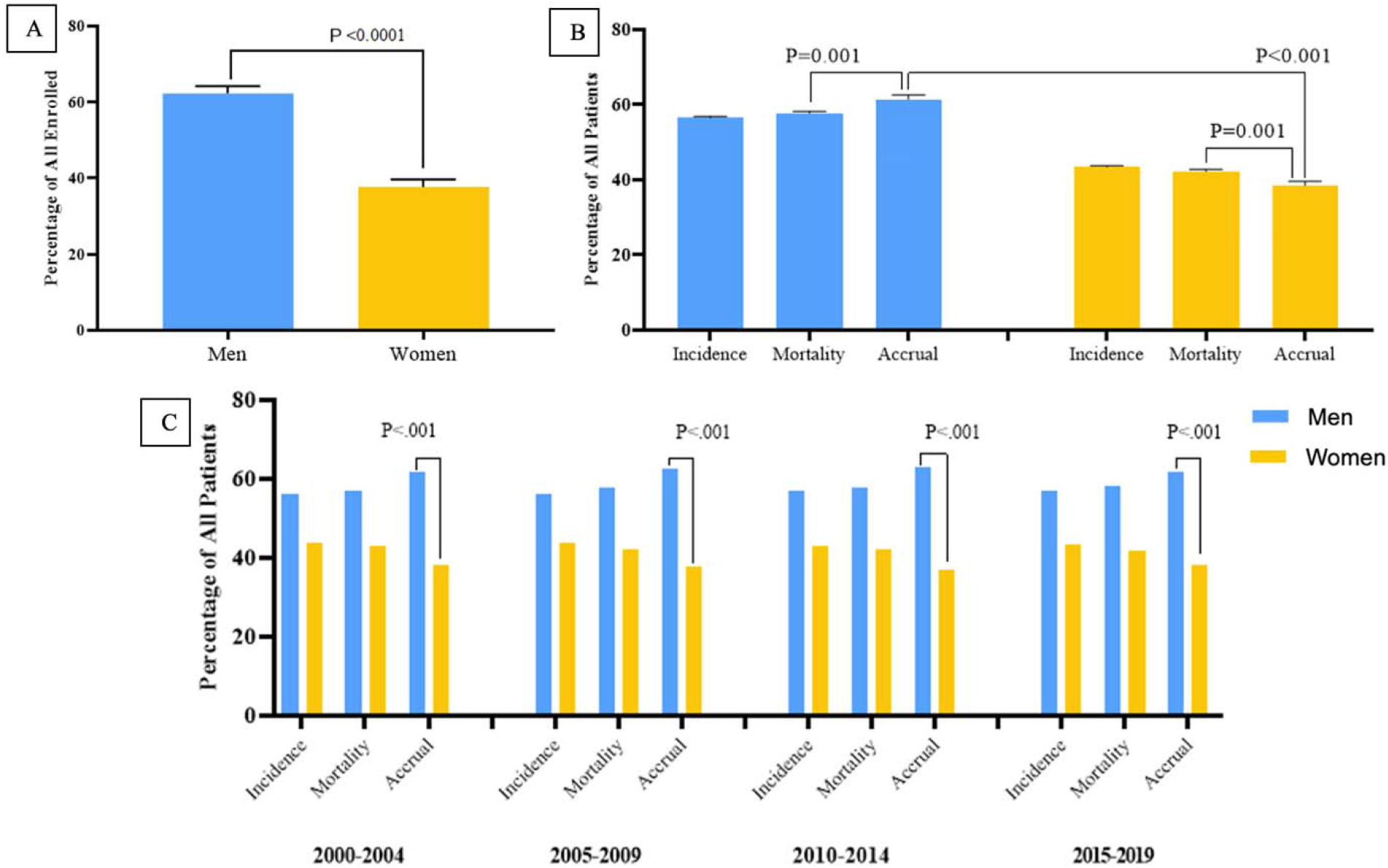
Proportions of Men and Women Enrolled in a Clinical Trial 2000-2019, Compared to Incidence and Mortality Burden. A. Clinical trial accrual proportions in men and women over the 20-year period, 2000-2019. Men represented 62.3% of accrued participants, women 37.7% (p<0.0001) B. Proportions of accrued participants as compared to disease incidence and mortality. Men were disproportionately accrued compared to their disease burden (p=0.001), and women were under-accrued compared to their disease burden (p=0.001). C. Five-year trends from 2000 to 2019 show consistently significant results across the time period. *Data Source: Incidence - CBTRUS: Data provided by CDC’s National Program of Cancer Registries and NCI’s Surveillance, Epidemiology and End Results Program, 2000-2017, Mortality - Incidence-Based Mortality SEER Research Data, (2000-2017), Accrual - Systematic review of the literature published of clinical (Phase I-IV) trials of adult gliomas (2000 – 2019).

Of the studies reporting sex over the 20-year period, men accounted for an average of 62.5% (n=26,237) of accrued participants. Regarding accrual by race, White participants accounted for 85.7% (n=10,806), Black/African Americans 2.0% (n=256), Asian/Pacific Islanders 1.3% (n=158), American Indian/Alaska Native 0.2% (n=20). Hispanic/Latino participants accounted for 1.9% (n=239) of reported participants in the studies that included data on ethnicity. Collectively, participants belonging to an NIH-designated minority group accounted for 5.3% (n=663) of the total reported participants included from studies with documented race/ethnicity (Table 1). An evaluation of the proportions (percentages) of each demographic group within each clinical trial revealed that on average articles reported sample populations that were 62.3% Male (standard deviation [SD]: 1.9%), 91.7% White (SD: 2.9%) and 5.9% Minority (SD: 3.4%). The Minority group was comprised of 2.6% Black/African American (SD: 2.2%), 1.5% Asian/Pacific Islander (SD:1.3%), 1.7% Hispanic-Latino (SD: 2.1%), 0.1% American Indian/Alaska Native (SD: 0.1%)(Table 1). Year-by-year proportions of each sex, race, and ethnicity group can be found in the Supplemental Figure 2.

**Figure 2:**
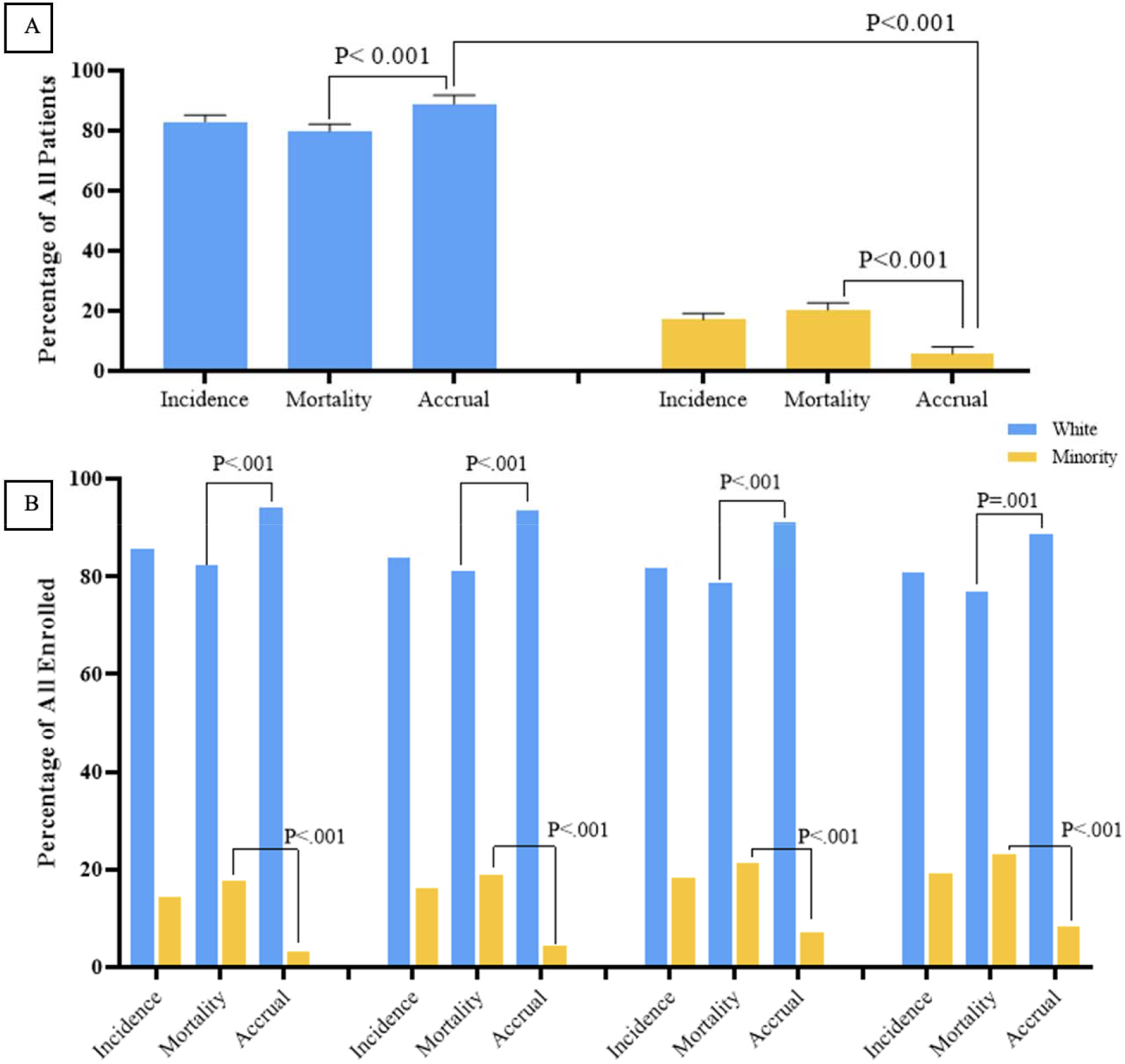
Patient Proportions for Incidence, Mortality, and Accrual According to Minority* Identity, 20-year Average and Five-Year Trends 2000-2019. A. Patient proportions for incidence, mortality, and accrual according to Minority* identity, 2000-2019. White participants represented 91.7% of accrued participants, Minority 5.9% (p<0.001). White patients were disproportionately accrued compared to their disease burden (p<0.001), and Minority patients were under-accrued compared to their disease burden (p<0.001). B. Five-Year Trends from 2000 to 2019 show consistently significant results across the time period. *Data Source: Incidence - CBTRUS: Data provided by CDC’s National Program of Cancer Registries and NCI’s Surveillance, Epidemiology and End Results Program, 2000-2017, Mortality - Incidence-Based Mortality SEER Research Data, (2000-2017), Accrual - Systematic review of the literature published of clinical (Phase I-IV) trials of adult gliomas (2000 – 2019). **Minority status determined by identification with an NIH-defined group.

### Disease incidence and mortality by demographic group

CBTRUS data included patients aged ≥20 years with newly diagnosed selected primary brain and central nervous (CNS) tumors that were either microscopically or radiographically confirmed for diagnosis from 2000 to 2017. The specific WHO ICD-O-3 histology codes included under each selected histology for analysis are included in Supplemental Table 1. Incidence data included a total of 257,663 incident-cases, of these 214,057 non-Hispanic White, 15,367 non-Hispanic Black, 22,145 Hispanic (all races), 4,879 Asian/Pacific Islander and 1,215 American Indian/Alaskan Native (Supplemental Table 2). Mortality data included a total of 45,765 deaths; 36,577 non-Hispanic Whites, 2,537 non-Hispanic Blacks, 4,468 Hispanic (all races), 2,046 Asian/Pacific Islanders and 137 American Indian/Alaskan Natives (Supplemental Table 3).

### Comparison of trial accrual and disease burden by demographic group

Over the 20-year period, men on average accounted for 62.3% of trial accrual, 55.9% of incident cases and 55.8% of disease mortality, while women accounted for 37.7% of trial accrual, 44.1% of incident cases and 44.2% of disease mortality (Figure 1.A). There was a statistically significant difference between accrual of men and women (DF: 23%, CI: 19%-26%, p<0.001) and between accrual and mortality for men (DF: 3.78 CI: 1.7%- 5.8%, p=.001, positive direction) and women (DF:−3.78 CI: −5.8%−11.7%, p=.001, negative direction) (Figure 1.B), which remained significant across 5-year trends (Figure 1.C). White participants accounted for 91.7% of trial accrual, 83.1% of incident cases, and 79.9% of deaths. The minority group at large, accounted for 5.9% of accrual, 16.9% of incident cases, and 20.1% of deaths. There was a statistically significant difference in accrual of Whites and Minorities (DF: 84% (CI: 77%-92%, p=<.001)(Figure 2.A.). There was a statistically significant difference between accrual and mortality for White (DF: 9% CI: 7%-11%, p<0.001, positive direction), the Minority group (DF:16% CI: 14%-18%, p<0.001) which were consistent across 5-year trends (Figure 2.B). Breakdown by race and ethnicity showed Black/African American accounted for 3.3% of trial accrual, 6.0% of incident cases, and 5.5% of mortalities; Hispanic/Latino: 1.8% of trial accrual, 8.6% of incidence, 9.8% of deaths, Asian/Pacific-Islander: 1.3% of trial accrual, 1.9% of incident cases, and 4.5% of deaths, American Indian/Alaska-Native: .06% of trial accrual, .5% of incident cases and .3% of deaths (accrual proportions in Figure 3). All minority racial/ethnic backgrounds showed significant under-accrual compared to their mortality burden; Black (DF: 3.1, CI: 1.5%-4.6%, p=.008), Hispanic/Latino (DF: 8.5%, CI: 7.6%-9.5%, p<.001), Asian (DF: 3.7%, CI: 3.1%-4.2%, p<.001), and Native (DF:0.3%, CI: 0.2%-0.5%, p=.008) (Figure 4).

**Figure 3:**
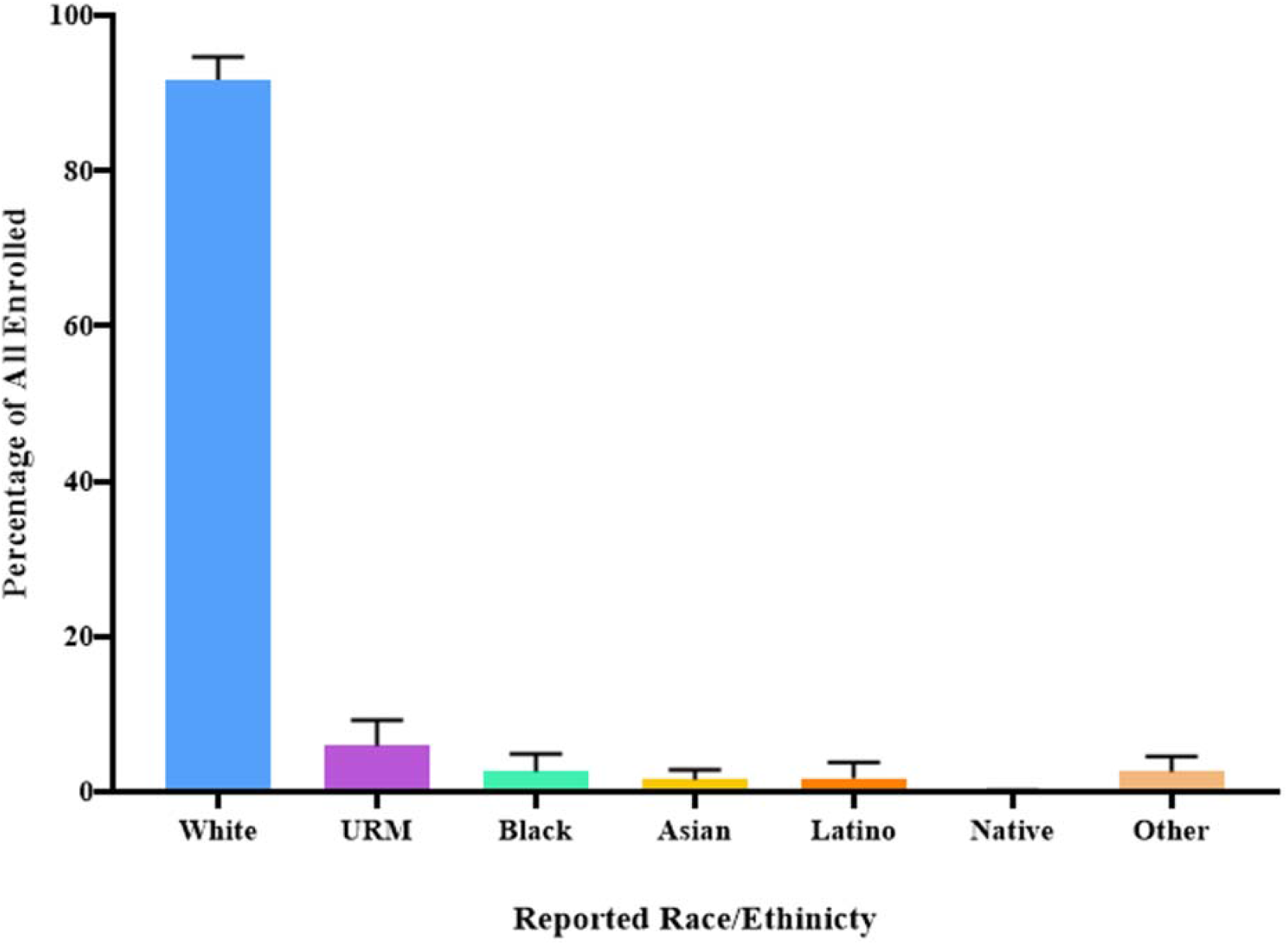
Clinical Trial Enrollment by Race and Ethnicity in Articles Published 2000-2019. White participants, on average, accounted for 91.7% of enrolled participants over the 20-year period. Under-represented minorities as a group, accounted for 5.9% of enrolled participants (Black/African American: 2.6%, Asian/Pacific Islander: 1.5%, Hispanic/Latino: 1.7%, Native American/Alaska Native: 0.1%) Abbreviations: URM: Under-represented minority

**Figure 4:**
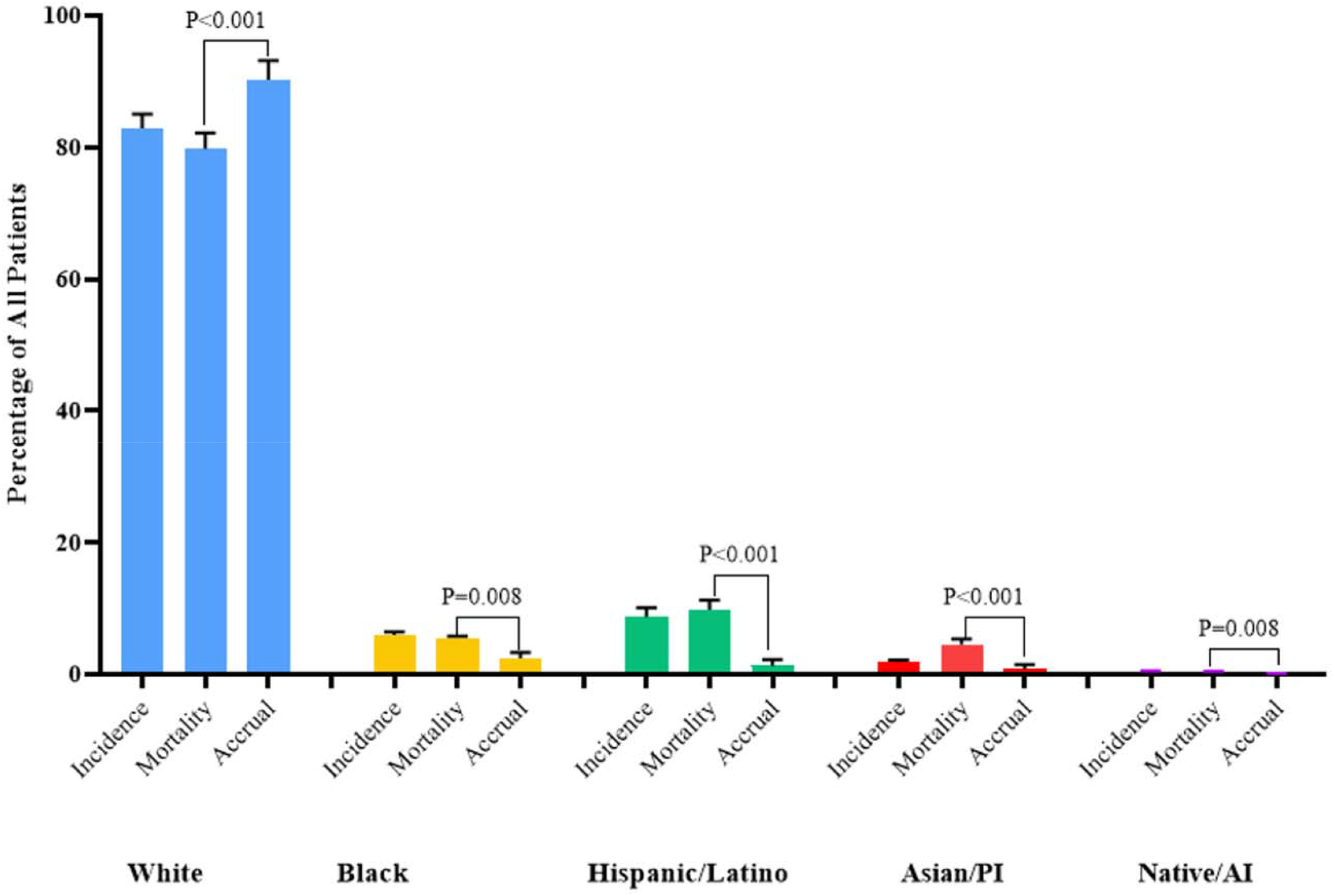
Patient Proportions for Incidence, Mortality, and Accrual According to Race and Ethnicity, 2000-2019. Over the 20-year period, White patients were disproportionately-accrued compared to their mortality burden 114% of expected (p<0.001), their minority counterparts were significantly under-accrued; Black/African-American 47% of expected (p=0.008), Hispanic/Latino 17% of expected (p<0.001), Asian/Pacific Islander 33% of expected (p<0.001) and Native American/Alaska Native 20% of expected (p=0.008). Abbreviations: PI: Pacific Islander, AI: Alaska Native *Data Source: Incidence - CBTRUS: Data provided by CDC’s National Program of Cancer Registries and NCI’s Surveillance, Epidemiology and End Results Program, 2000-2017, Mortality - Incidence-Based Mortality SEER Research Data, (2000-2017), Accrual - Systematic review of the literature published of clinical (Phase I-IV) trials of adult gliomas (2000 – 2019).

We computed a measure of enrollment by demographic group using Enrollment Incidence Ratio (EIR) and Enrollment Mortality Ratio (EMR) (defined in Supplemental Methods). The overall EIR was 1.10 in men, 0.89 in women, 1.1 in White patients, and 0.35 in Minority patients (0.44 in Black, 0.211 in Hispanic/Latino, 0.78 in Asian/PI, 0.13 in Native/AI). By EIR, men had 1.25 greater odds of enrollment compared to women, while White patients had 3.95 greater odds of enrollment compared to their Minority counterparts. The overall EMR was 1.08 in men, 0.89 in women, 1.15 in White patients, and 0.29 in Minority patients (0.47 in Black, 0.17 in Hispanic/Latino, 0.33 in Asian/PI, 0.20 in Native/AI). By EMR, men had 1.20 greater odds of enrollment compared to women, while White patients had 3.76 greater odds of enrollment compared to their Minority counterparts.

### High-Accruing Studies

Seventeen studies were identified as upper tercile, in which minority recruitment met or exceeded 10% of enrollment. Within these studies, 55% of papers included a primary and/or senior author who self-identified as representing an NIH-defined minority group. These programs were in geographic areas with 60.5% minority population on average (2010-2016 Census Data). Their respective departments of Neurology and Neurosurgery were diverse, on average consisting of 43% minorities, exceeding the national average for U.S. medical school faculty diversity (28.5% minorities) and physician workforce diversity in neurology and neurosurgery combined (22.7% minorities; 24% in neurology & 22% in neurosurgery).

## Discussion

Within the U.S. significant differences in health outcomes exist between specific patient demographics. While health disparities are often interpreted as differences in outcomes between racial or ethnicity groups, disparities can exist across many dimensions, including gender, age, sexual orientation, socioeconomics, and disability status. The goal of this study was to analyze whether clinical trial accrual within neuro-oncology differentiates from patterns of disease-specific incidence and mortality during the 20-year period following the NIH Revitalization Act. The present study demonstrates that White males remain disproportionately represented in clinical trials for adult neuro-epithelial CNS tumors despite having a slightly higher rate of incidence compared to women. Minority patients are diagnosed with neuro-epithelial tumors at a lower rate compared with White patients; however, they suffer from a striking underrepresentation in trial accrual based on incidence and mortality.

Clinical trial accrual for women and minorities remains low 27 years since the implementation of the NIH Revitalization Act. This study demonstrates that not only does the disparity in clinical trial participation for CNS neuro-epithelial tumors remain significant for all minority groups, but the trajectory of accrual over time for underrepresented populations has not significantly improved. Between 2000-2019. clinical trial race and gender reporting was poor and when reported, accrual did not meet the burden of mortality or incidence for Black/African-Americans, Hispanic/Latinos, Asian/Pacific-Islanders, or American-Indian/Alaska Natives. Furthermore, there has been minimal upward improvement in accrual over the last 10 years. With respect to gender, the inclusion of women in clinical trials has seen steady gains over the last two decades in the general cancer population. Within neuro-oncology however gender enrollment disparities have remained sluggish when compared with men^3^. Furthermore, only a fraction of clinical trials reported race and ethnicity data, and among those that did report, there were major inconsistencies in the manner in which demographic information was presented. Studies that were able to recruit more than 10% minorities, appeared to have diverse department faculty, currently, and located in geographic areas with higher proportions of minorities.

Disparities persist in delivery of standard of care and experimental cancer directed therapies in neuro-oncology. For example, non-Hispanic Blacks and Hispanics remain less likely to receive chemoradiation when compared with non-Hispanic Whites^10^. A recent review of NCI-sponsored clinical trials found persistent and significant under-reporting and under-enrollment of minorities in cancer studies. The authors found that less than 2% of clinical trials had a primary purpose of investigating cancer in minority populations^3^. There are a number of obstacles to be confronted in efforts to improve accrual of minorities in clinical trials. The conceptual framework first conceived by Ford, et. al and later adapted by Napoles et. al, provides an example of the complexity in understanding patient-level decision making in considering participation in a clinical trial. There are numerous barriers including those of awareness, knowledge, opportunity, all of which remain formidable challenges in clinical trial design and implementation, particularly for diverse populations.

Transparency in reporting results of clinical trials is needed in order to accomplish any meaningful change in accrual in neuro-oncology,, particularly for subgroup analyses by race and ethnicity. Collective, purposeful efforts are needed to standardize the manner in which clinical research data is collected and reported. Achieving race, ethnic, and gender equity in scientific research is not just a moral cause. Diversity in clinical trials, and clinical research at large, is paramount to strengthening our ability to affirm validity of findings that advance both mechanistic understanding of disease and medical interventions. Appropriate inclusion of all affected demographic groups, at all levels of investigation, is central to the path of both health justice and precision medicine.

We acknowledge that the impact of race/ethnicity as a social determinant of health does not exist in a vacuum and is profoundly impacted by and alongside various socioeconomic, environmental, and structural factors, particularly in cancer research and treatment. For example, given that only 20% of published articles in neurooncology included detailed breakdowns of race and/or ethnicity of participants, when considering the accrual of Hispanic/Latino participants prior to the introduction of formal accounting for ethnicity, we cannot determine with certainty, the articles that did not report specific minority groups, truly did not enroll a considerable number of Hispanic/Latino participants. For this reason, specific clinical trial reporting standards should be observed. In order to address the central issues of healthcare disparities in cancer research, we must begin with both accurate and precise data collection along with continued emphasis on recruiting and retaining diverse populations that meet the needs of demographic-specific disease burden.

## Conclusion

Despite the NIH mandate, introduced almost three decades ago, a clear disparity remains between the accrual of minorities and women in clinical trials for adult neuro-epithelial tumors as compared to their respective disease burden and representation in the U.S. population. Although the gap in enrollment for women has improved dramatically in the general cancer population it remains stagnant in neuro-oncology trials. Representation of minorities in clinical trials remains significantly below disease burden. The upward trend in accrual across minority groups, however, is a signal for continued efforts towards interventions for improving participation and standardizing reporting. The quality of scientific research and the knowledge base in neuro-oncology can only be strengthened by increased diversity. With improved representation comes immense potential to improve the clinical outcomes for groups that often bear disproportionate burden.

## Data Availability

All data used in this study are presented in the manuscript, supplemental data, and are available through review of published clinical trials on PubMed and/o ClinicalTrials.gov

## Abbreviations

DF: mean of differences
CI: confidence interval
CNS: central nervous system

