## Supplemental Methods & Data for "A Population Study of Clinical Trial Accrual for Women and Minorities in Neuro-Oncology Following the NIH Revitalization Act"

**Supplementary Methods**

**Accrual Data Extracted from Published Papers**

Patient demographic characteristics, to include gender, race, and ethnicity were collected from all eligible articles. Sixteen articles were excluded because they did not report sex or race/ethnicity. “Minority” status was defined as belonging to any of the NIH-designated underserved minority groups in clinical research, to include Black/Africa-American, Asian/Native Hawaiian/Pacific Islander, American-Indian/Alaska Native, or of Hispanic/Latino ethnicity. It is important to note that Hispanic/Latino includes individual that identify as Hispanic White and Hispanic Black. Each article’s published text and supplemental materials were scanned for participant demographics. If not explicitly reported in the article itself and the trial NCT# was available, a subsequent search was conducted on ClinicalTrials.gov site to gather this information. For each demographic factor, absolute counts and proportions of total reported were collected by calendar year, and average percentages were calculated.

Enrollment to incidence ratio and enrollment to mortality ratios were calculated using the enrolled proportion for each demographic group as the numerator to the incident- and mortality-based disease burden, respectively as the denominator.

**Incidence and Mortality Rate for Primary Malignant Brain and CNS tumors by Race and Ethnicity**

**Selection criteria for the study:** Patients aged ≥20 years with newly diagnosed selected neuro-epithelial tumors of the brain and central nervous tumors that were either microscopically or radiographically confirmed for diagnosis years 2000 to 2017 were included. The specific WHO ICD-O-3 histology codes included under each histology are included in Supplemental Table 1.

**Data Source and Statistical Methods:**

*Data sources*

The Central Brain Tumor Registry of the United States (CBTRUS) database was used to estimate the age-adjusted incidence rates.^11^ *CBTRUS Incidence Data:* Central Brain Tumor Registry of the United States SEER*Stat Database. CDC National Program of Cancer Registries and NCI Surveillance, Epidemiology and End Results Incidence Data,  2019 submission (2000-2017)^12^. *SEER Incidence-Based Mortality Data:*  SEER Program (www.seer.cancer.gov) SEER*Stat Database: Incidence-Based Mortality - SEER Research Data, 18 Registries, Nov 2019 Sub (2000-2017) - Linked to County Attributes - Time Dependent (1990-2017) Income/Rurality, 1969-2018 Counties, National Cancer Institute, DCCPS, Surveillance Research Program. Released April 2020, based on the 2019 submission^13,14^.

CBTRUS is a population-based site-specific registry that obtains the latest available data on all newly diagnosed primary brain and other CNS tumors from the Centers for Disease Control and Prevention’s (CDC) National Program of Cancer Registries (NPCR), and the National Cancer Institute’s (NCI) Surveillance, Epidemiology, and End Results (SEER) program. These data include de-identified incidence data from 52 Central Cancer Registries (48 NPCR and 4 SEER [SEER data available until year 2016 only]) for malignant and non-malignant (benign and uncertain behaviors) primary brain and other CNS tumors. Average annual age-adjusted incidence rates (AAAIR) and 95% confidence intervals (95% CI) were estimated per 100,000 population, based on one-year age groupings and standardized to the 2000 US standard population (Supplemental Table 2).^13^

Incidence-based age-adjusted mortality rates (AAMR), estimated mortality using population-level cancer registry data, were calculated using the data from 18 central cancer registries included in the SEER 18.^14^ These registries represent 28% of the US population and are a subset of those registries included in the overall CBTRUS analytic dataset**.** Once a cancer has been reported to the SEER Program, the case is followed to monitor vital status. As a result, a cause-specific mortality rate among those persons diagnosed with cancer and reported to the SEER Program can be calculated. The numerator for the “incidence-file-based mortality rate” consists of the number of primary malignant brain and other CNS tumors deaths among those persons with a primary malignant brain and other CNS tumors diagnosis reported to the cancer registry. The denominator for this rate is the population at risk at the time of the deaths. Caution must be used in interpreting these results, as they can be affected by factors, such as reporting delay and lead-time bias. Counts, rates and other relevant statistics were calculated using SEER*Stat 8.3.8.^15^ Rates are suppressed when counts are fewer than 20 within a cell.

**Supplementary Titles & Legends**

**Supplemental Figure 1. Study schema for selection and inclusion of articles in PubMed query.**

**Supplemental Figure 2: Year-by-Year Accrual Proportions by Sex, Race & Ethnicity 2000-2019**

*2002 data is absent on the Race/Ethnicity chart as none of the published studies in that year reported Race/Ethnicity of participants.

**Supplemental Table 1: Central Brain Tumor Registry of the United States (CBTRUS), Brain and Other Central Nervous System Tumor Selected Histology Groupings**

**Supplemental Table 2: Eighteen-Year Total, Average Annual Age-Adjusted Incidence Rates^a^ with 95% Confidence Intervals for Selected Primary Brain and Other Central Nervous System Tumors in Aged ≥20 years by Sex, CBTRUS: Data provided by CDC’s National Program of Cancer Registries and NCI’s Surveillance, Epidemiology and End Results Program, 2000-2017**

**Supplemental Table 3: Eighteen-Year Total, Average Annual Age-Adjusted Incidence Rates^a^ with 95% Confidence Intervals for Selected Primary Brain and Other Central Nervous System Tumor Histologies in Aged ≥20 years by Race and Ethnicity. (CBTRUS: Data provided by CDC’s National Program of Cancer Registries and NCI’s Surveillance, Epidemiology and End Results Program, 2000-2017).**

**Supplemental Table 4: Eighteen-Year Total and Average Annual Age-Adjusted Incidence-Based Mortality Rates^a^ with 95% Confidence Intervals for Selected Primary Brain and Other Central Nervous System Tumors in Aged ≥20 years by Sex, SEER, 2000-2017**

**Supplemental Table 5. Eighteen-Year Total and Average Annual Age-Adjusted Incidence-Based Mortality Rates^a^ with 95% Confidence Intervals for Selected Primary Brain and Other Central Nervous System Tumor Histologies in Aged ≥20 years by Race and Ethnicity (SEER Incidence-Based Mortality, 18 Registries, 2000-2017).**

**Supplemental Table 6: Five-Year Total, Average Annual Age-Adjusted Incidence Rates^a^ with 95% Confidence Intervals for Selected Primary Brain and Other Central Nervous System Tumors^b^ in Aged ≥20 years by Sex, CBTRUS: Data provided by CDC’s National Program of Cancer Registries and NCI’s Surveillance, Epidemiology and End Results Program, 2000-2017**

**Supplemental Table 7: Five-Year Total, Average Annual Age-Adjusted Incidence Rates^a^ with 95% Confidence Intervals for Selected Primary Brain and Other Central Nervous System Tumors^b^ in Aged ≥20 years by Race/Ethnicity, CBTRUS: Data provided by CDC’s National Program of Cancer Registries and NCI’s Surveillance, Epidemiology and End Results Program, 2000-2017**

**Supplemental Table 8: Five-Year Total, Average Annual Age-Adjusted Incidence-Based Mortality Rates^a^ with 95% Confidence Intervals for Selected Primary Brain and Other Central Nervous System Tumors^b^ in Aged ≥20 years by Sex, SEER, 2000-2017**

**Supplemental Table 9: Five-Year Total, Average Annual Age-Adjusted Incidence-Based Mortality Rates^a^ with 95% Confidence Intervals for Selected Primary Brain and Other Central Nervous System Tumors^b^ in Aged ≥20 years by Race/Ethnicity, SEER, 2000-2017**

**Supplemental Figure 3: Five-Year Grouped Trends of Incidence, Mortality, and Accrual by Reported Sex 2000-2019.**

*Data Source: Incidence - CBTRUS: Data provided by CDC’s National Program of Cancer Registries and NCI’s Surveillance, Epidemiology and End Results Program, 2000-2017, Mortality - Incidence-Based Mortality SEER Research Data, (2000-2017), Accrual - Systematic review of the literature published of clinical (Phase I-IV) trials of adult gliomas (2000 – 2019).

**Supplemental Figure 4: Five-Year Grouped Trend Data for Incidence, Mortality and Accrual by Minority* Status 2000-2019**

Minority status determined by identification with an NIH-defined group.

*Data Source: Incidence - CBTRUS: Data provided by CDC’s National Program of Cancer Registries and NCI’s Surveillance, Epidemiology and End Results Program, 2000-2017, Mortality - Incidence-Based Mortality SEER Research Data, (2000-2017), Accrual - Systematic review of the literature published of clinical (Phase I-IV) trials of adult gliomas (2000 – 2019).

**Supplemental Figure 5: Five-Year Grouped Trend Data for Incidence, Mortality and Accrual by Race and Ethnicity 2000-2019**

*Data Source: Incidence - CBTRUS: Data provided by CDC’s National Program of Cancer Registries and NCI’s Surveillance, Epidemiology and End Results Program, 2000-2017, Mortality - Incidence-Based Mortality SEER Research Data, (2000-2017), Accrual - Systematic review of the literature published of clinical (Phase I-IV) trials of adult gliomas (2000 – 2019).

**Supplemental Figure 1. Study schema for selection and inclusion of articles in PubMed query.**

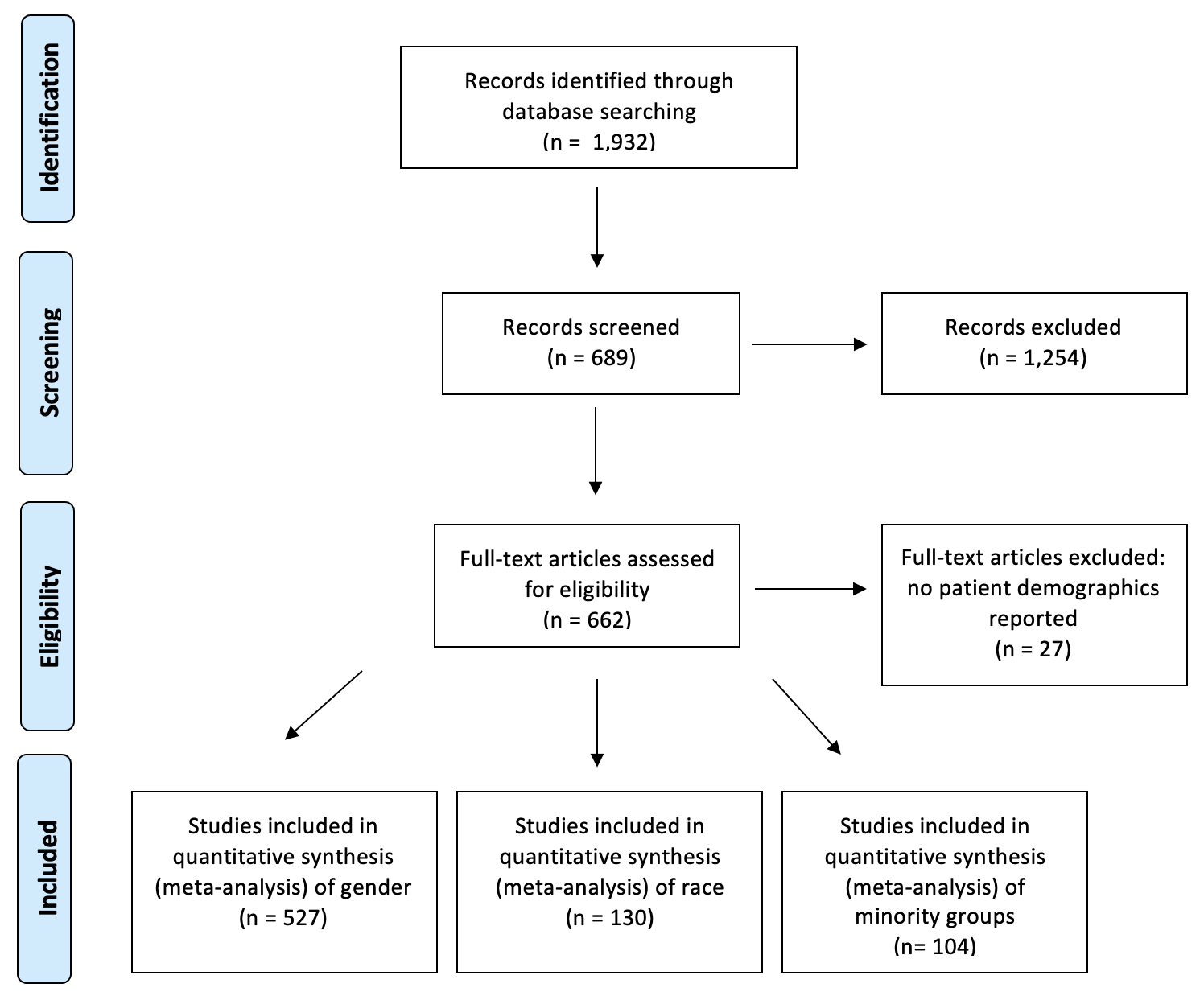

**Supplemental Figure 2: Year-by-Year Accrual Proportions by Sex, Race & Ethnicity 2000-2019**

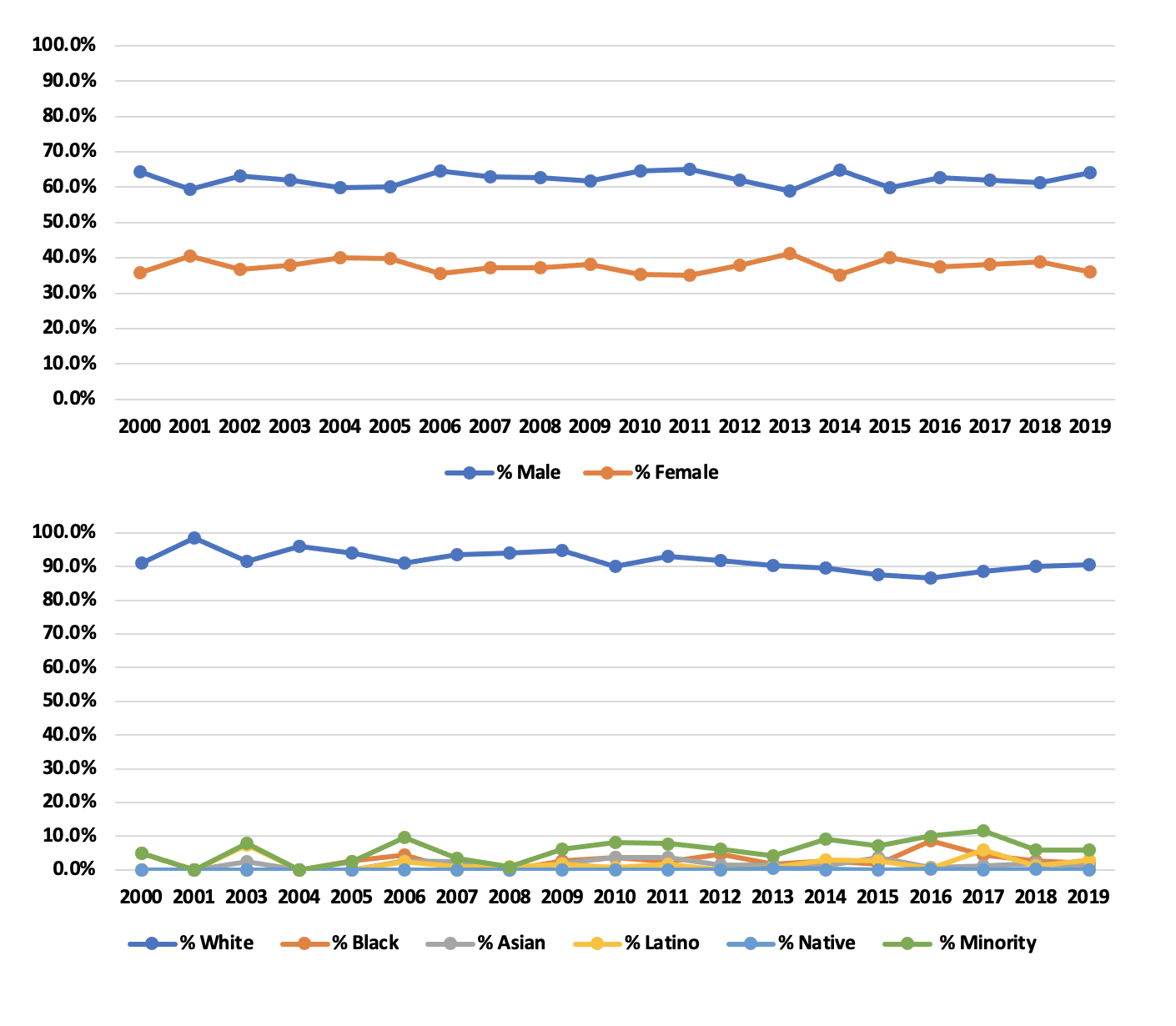

**Supplemental Table 1: Central Brain Tumor Registry of the United States (CBTRUS), Brain and Other Central Nervous System Tumor Selected Histology Groupings.**

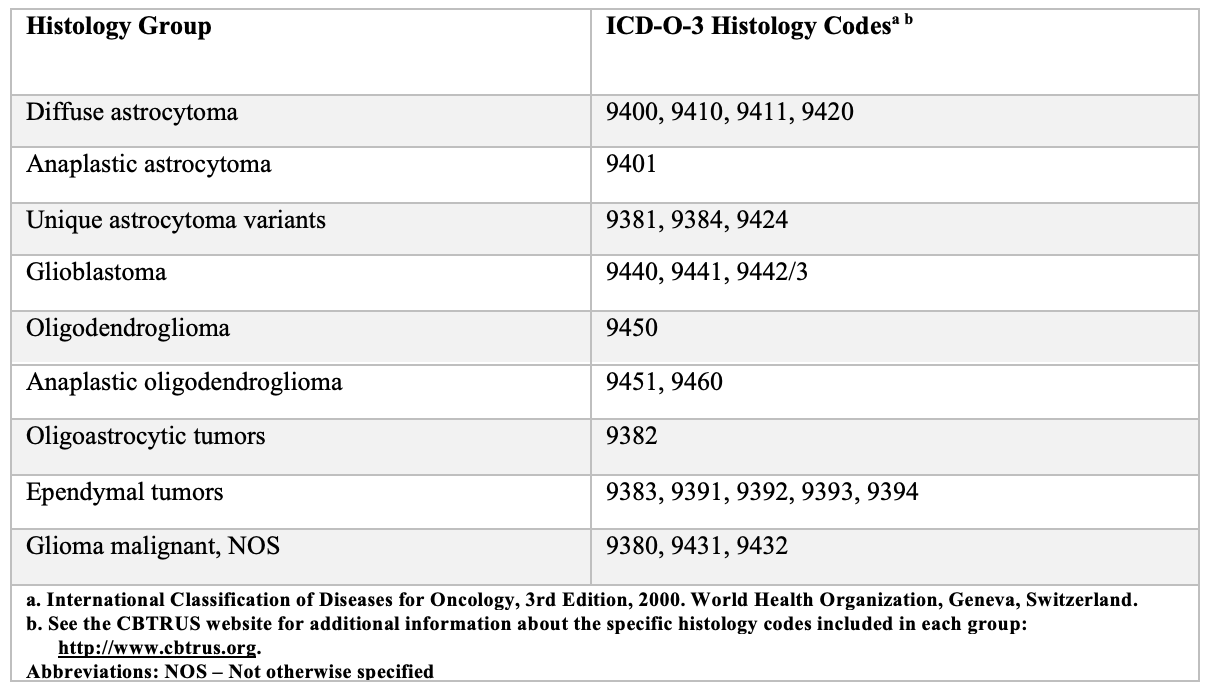

**Supplemental Table 2: Eighteen-Year Total, Average Annual Age-Adjusted Incidence Rates^a^ with 95% Confidence Intervals for Selected Primary Brain and Other Central Nervous System Tumors in Aged ≥20 years by Sex, CBTRUS: Data provided by CDC’s National Program of Cancer Registries and NCI’s Surveillance, Epidemiology and End Results Program, 2000-2017**

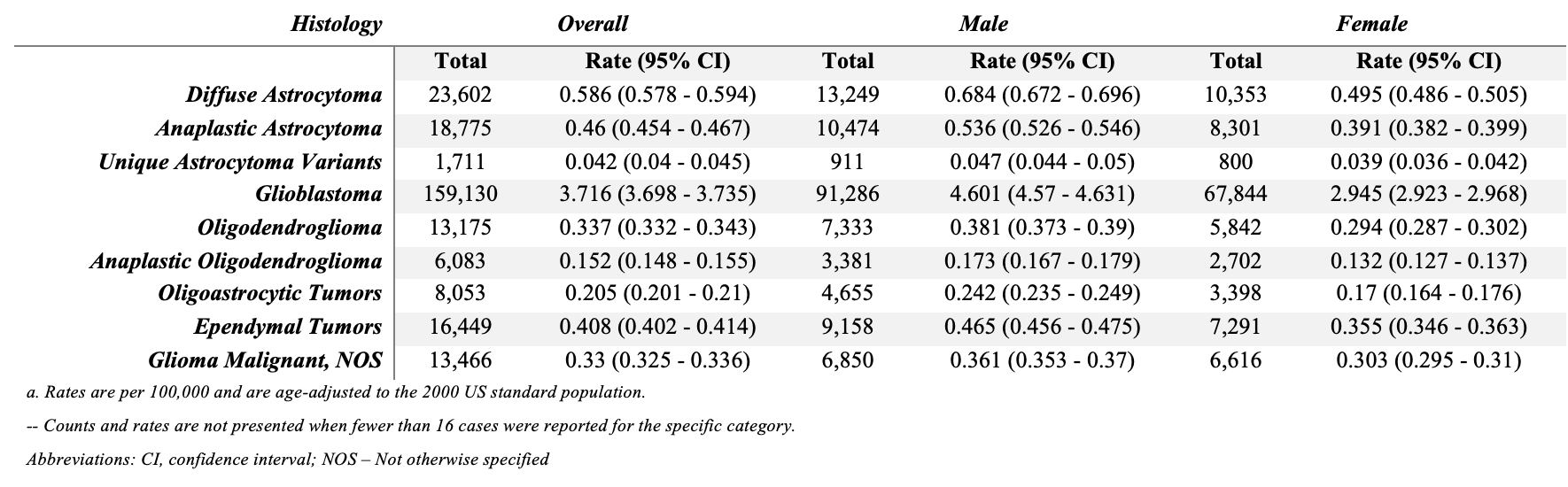

**Supplemental Table 3: Eighteen-Year Total, Average Annual Age-Adjusted Incidence Rates^a^ with 95% Confidence Intervals for Selected Primary Brain and Other Central Nervous System Tumor Histologies in Aged ≥20 years by Race and Ethnicity. (CBTRUS: Data provided by CDC’s National Program of Cancer Registries and NCI’s Surveillance, Epidemiology and End Results Program, 2000-2017).**

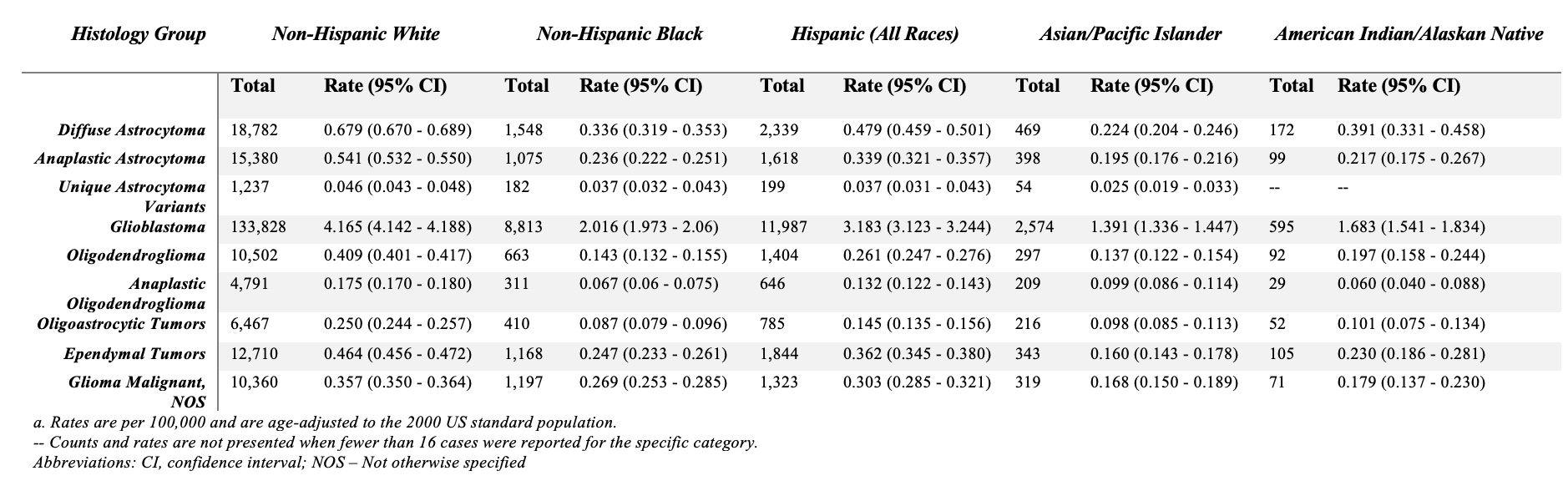

**Supplemental Table 4: Eighteen-Year Total and Average Annual Age-Adjusted Incidence-Based Mortality Rates^a^ with 95% Confidence Intervals for Selected Primary Brain and Other Central Nervous System Tumors in Aged ≥20 years by Sex, SEER, 2000-2017**

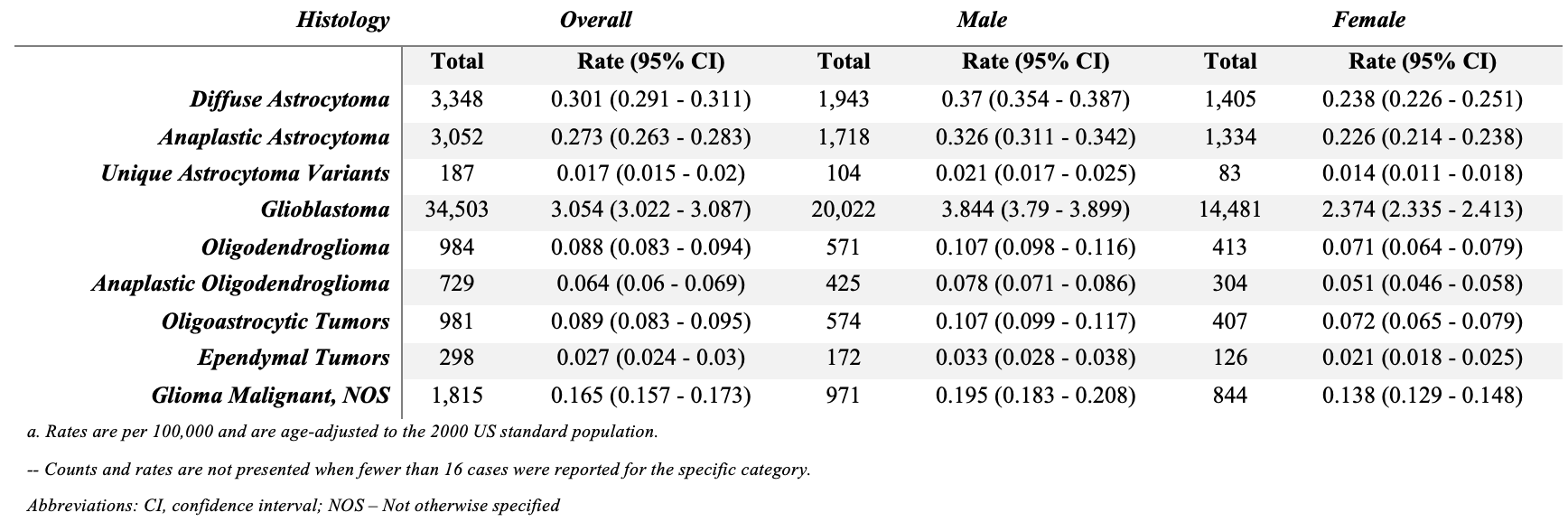

**Supplemental Table 5. Eighteen-Year Total and Average Annual Age-Adjusted Incidence-Based Mortality Rates^a^ with 95% Confidence Intervals for Selected Primary Brain and Other Central Nervous System Tumor Histologies in Aged ≥20 years by Race and Ethnicity (SEER Incidence-Based Mortality, 18 Registries, 2000-2017).**
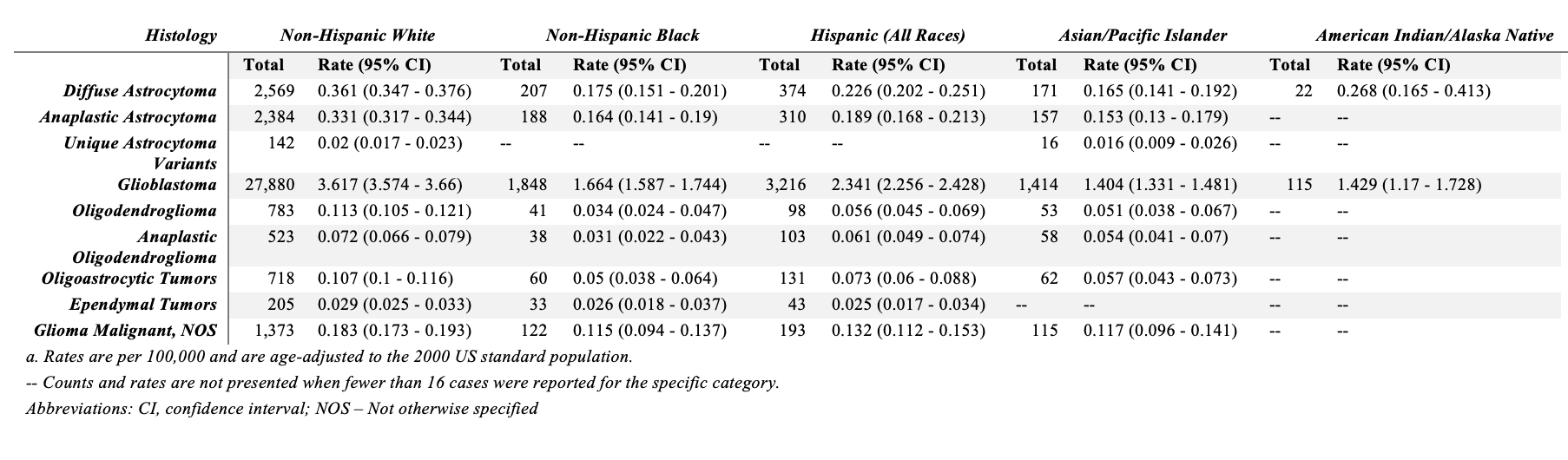

**Supplemental Table 6: Five-Year Total, Average Annual Age-Adjusted Incidence Rates^a^ with 95% Confidence Intervals for Selected Primary Brain and Other Central Nervous System Tumors^b^ in Aged ≥20 years by Sex, CBTRUS: Data provided by CDC’s National Program of Cancer Registries and NCI’s Surveillance, Epidemiology and End Results Program, 2000-2017**

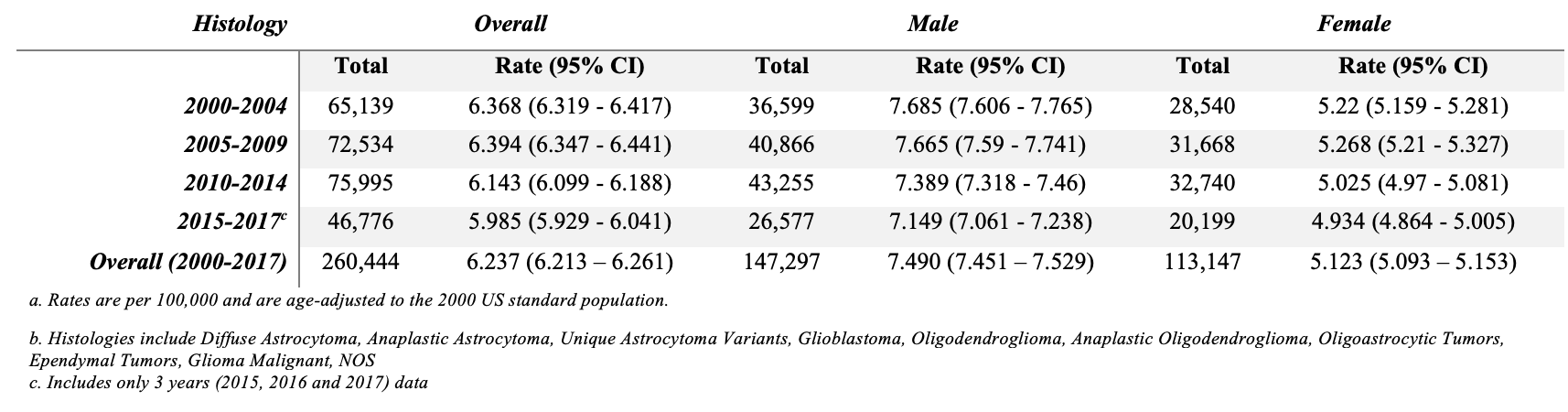

**Supplemental Table 7: Five-Year Total, Average Annual Age-Adjusted Incidence Rates^a^ with 95% Confidence Intervals for Selected Primary Brain and Other Central Nervous System Tumors^b^ in Aged ≥20 years by Race/Ethnicity, CBTRUS: Data provided by CDC’s National Program of Cancer Registries and NCI’s Surveillance, Epidemiology and End Results Program, 2000-2017**

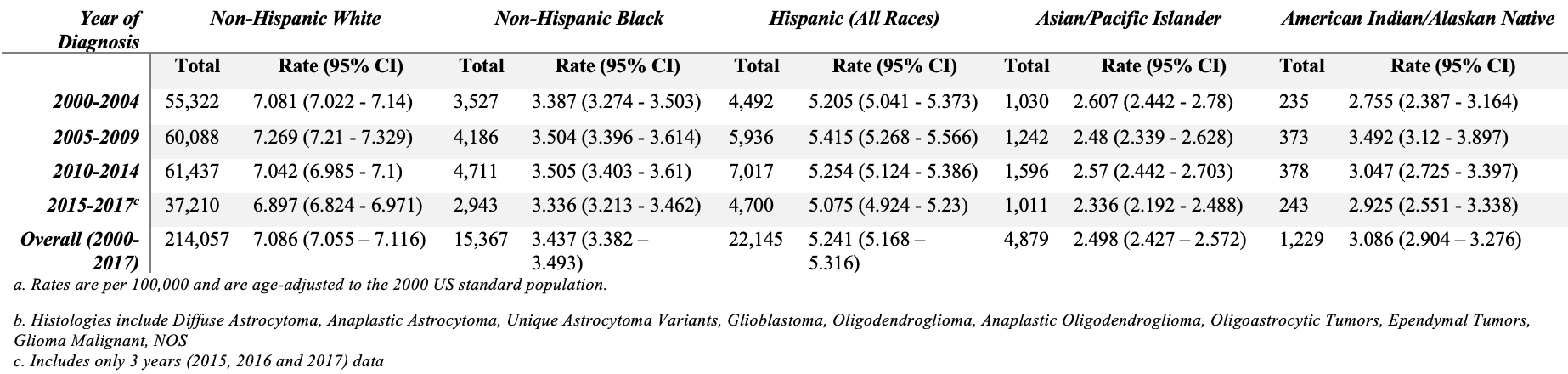

**Supplemental Table 8: Five-Year Total, Average Annual Age-Adjusted Incidence-Based Mortality Rates^a^ with 95% Confidence Intervals for Selected Primary Brain and Other Central Nervous System Tumors^b^ in Aged ≥20 years by Sex, SEER, 2000-2017**

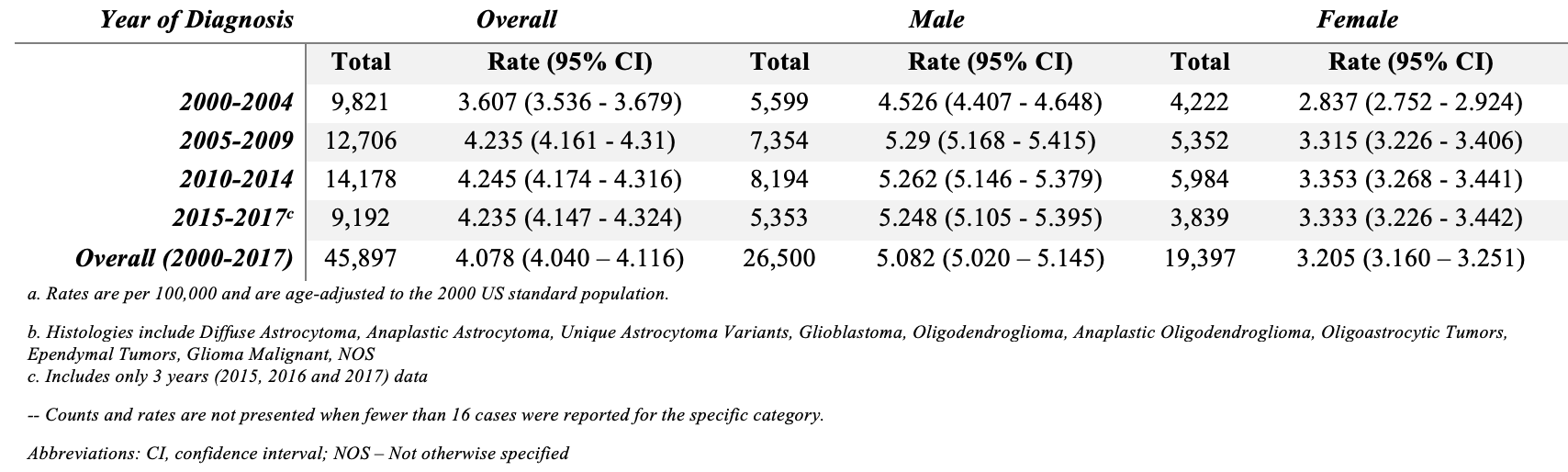

**Supplemental Table 9: Five-Year Total, Average Annual Age-Adjusted Incidence-Based Mortality Rates^a^ with 95% Confidence Intervals for Selected Primary Brain and Other Central Nervous System Tumors^b^ in Aged ≥20 years by Race/Ethnicity, SEER, 2000-2017**

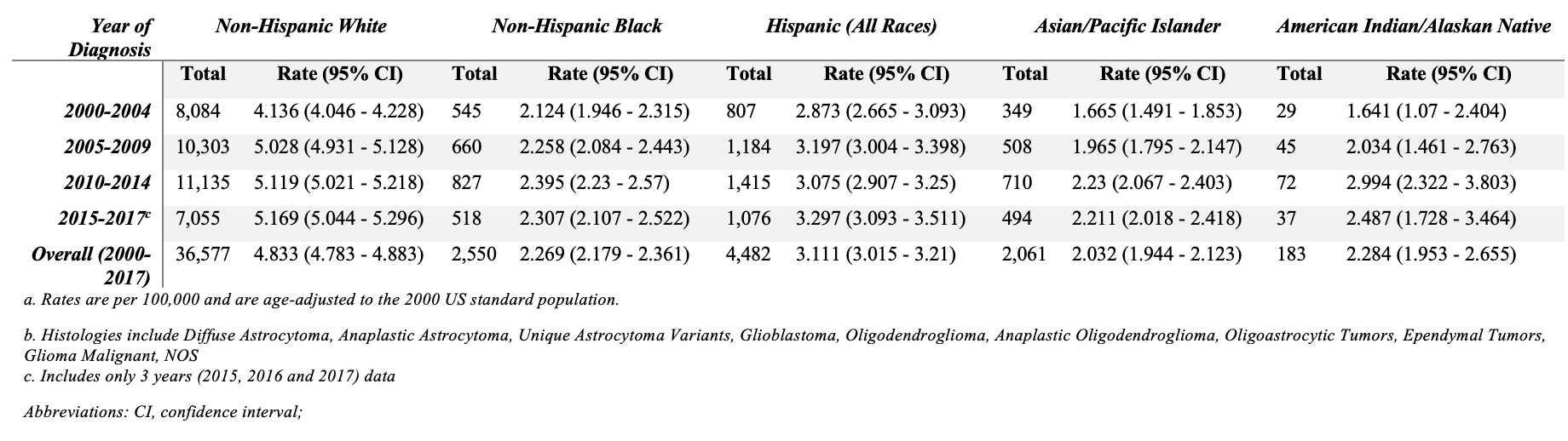

**Supplemental Figure 3: Five-Year Grouped Trends of Incidence, Mortality, and Accrual by Reported Sex 2000-2019.**

Percentage of All Patients

|  | 2000-2004 | 2005-2009 | 2010-2014 | 2015-2019 |
| --- | --- | --- | --- | --- |
| Men Incidence | 56.186% | 56.3405% | 56.9182% | 56.8176% |
| Men Mortality | 57.0105% | 57.8782% | 57.7938% | 58.2354% |
| Men Accrual | 61.7% | 62.4% | 63.1% | 61.9% |
| Women Incidence | 43.814% | 43.6595% | 43.0818% | 43.1824% |
| Women Mortality | 42.9895% | 42.1218% | 42.2062% | 41.7646% |
| Women Accrual | 38.3% | 37.6% | 36.9% | 38.1% |

**Supplemental Figure 4: Five-Year Grouped Trend Data for Incidence, Mortality and Accrual by Minority* Status 2000-2019**

Percentage of All Patients

|  | 2000-2004 | 2005-2009 | 2010-2014 | 2015-2019 |
| --- | --- | --- | --- | --- |
| White Incidence | 85.6298% | 83.6589% | 81.7645% | 80.7036% |
| White Mortality | 82.3721% | 81.126% | 78.6426% | 76.8519% |
| White Accrual | 94.2% | 93.4% | 91% | 88.6% |
| Minority* Incidence | 14.3702% | 16.3411% | 18.2355% | 19.2964% |
| Minority* Mortality | 17.6279% | 18.874% | 21.3574% | 23.1481% |
| Minority* Accrual | 3.2% | 4.5% | 7% | 8.2% |
